# Clinical characteristics of children and young people hospitalised with covid-19 in the United Kingdom: prospective multicentre observational cohort study

**DOI:** 10.1101/2020.07.14.20153320

**Authors:** Olivia V Swann, Karl Holden, Lance Turtle, Louisa Pollock, Cameron J Fairfield, Thomas M Drake, Sohan Seth, Conor Egan, Hayley E Hardwick, Sophie Halpin, Michelle Girvan, Chloe Donohue, Mark Pritchard, Latifa B Patel, Shamez Ladhani, Louise Sigfrid, Ian P Sinha, Piero Olliaro, Jonathan S Nguyen-Van-Tam, Peter W Horby, Laura Merson, Gail Carson, Jake Dunning, Peter JM Openshaw, J Kenneth Baillie, Ewen M Harrison, Annemarie Docherty, Malcolm G Semple, on behalf of ISARIC4C Investigators

## Abstract

**Objective:** To characterise the clinical features of children and young people admitted to hospital with laboratory-confirmed severe acute respiratory syndrome coronavirus 2 (SARS-CoV-2) infection in the UK, and explore factors associated with admission to critical care, mortality, and development of multisystem inflammatory syndrome in children and adolescents temporarily related to covid-19 (MIS-C).

**Design:** Prospective observational cohort study with rapid data gathering and near real time analysis.

**Setting:** 260 acute care hospitals in England, Wales, and Scotland between 17th January and 5^th^ June 2020, with a minimal follow-up time of two weeks (to 19^th^ June 2020).

**Participants:** 451 children and young people aged less than 19 years admitted to 116 hospitals and enrolled into the International Severe Acute Respiratory and emergency Infections Consortium (ISARIC) WHO Clinical Characterisation Protocol UK study with laboratory-confirmed SARS-CoV-2.

**Main Outcome Measures:** Admission to critical care (high dependency or intensive care), in-hospital mortality, or meeting the WHO preliminary case definition for MIS-C.

**Results:** Median age was 3.9 years [interquartile range (IQR) 0.3-12.9 years], 36% (162/451) were under 12 months old, and 57% (256/450) were male. 56% (224/401) were White, 12% (49/401) South Asian and 10% (40/401) Black. 43% (195/451) had at least one recorded comorbidity. A muco-enteric cluster of symptoms was identified, closely mirroring the WHO MIS-C criteria.

17% of children (72/431) were admitted to critical care. On multivariable analysis this was associated with age under one month odds ratio 5.05 (95% confidence interval 1.69 to 15.72, p=0.004), age 10 to 14 years OR 3.11 (1.21 to 8.55, p=0.022) and Black ethnicity OR 3.02 (1.30 to 6.84, p=0.008). Three young people died (0.7 %, 3/451) aged 16 to 19 years, all of whom had profound comorbidity.

Twelve percent of children (36/303) met the WHO MIS-C criteria, with the first patient developing symptoms in mid-March. Those meeting MIS-C criteria were older, (median age 10.8 years ([IQR 8.4-14.1] vs 2.0 [0.2-12.6]), p<0.001) and more likely to be of non-White ethnicity (70% (23/33) vs 43% (101/237), p=0.005). Children with MIS-C were four times more likely to be admitted to critical care (61% (22/36) vs 15% (40/267, p<0.001). In addition to the WHO criteria, children with MIS-C were more likely to present with headache (45% (13/29) vs 11% (19/171), p<0.001), myalgia (39% (11/28) vs 7% (12/170), p<0.001), sore throat (37% (10/27) vs (13% (24/183, p = 0.004) and fatigue (57% (17/30) vs 31% (60/192), p =0.012) than children who did not and to have a platelet count of less than 150 ×10^9^/L (30% (10/33) vs 10% (24/232), p=0.004).

**Conclusions:** Our data confirms less severe covid-19 in children and young people than in adults and we provide additional evidence for refining the MIS-C case definition. The identification of a muco-enteric symptom cluster also raises the suggestion that MIS-C is the severe end of a spectrum of disease.

**Study registration:** ISRCTN66726260

## Introduction

Children and young people comprise only 1-2% of covid-19 cases worldwide [1–3]. In contrast to other respiratory viruses, children appear to have a lower risk of infection than adults [4] and the vast majority of reported infections in children are reported as mild or asymptomatic, with few recorded childhood fatalities attributed to covid-19 [2,5–7]. Initial reports from China showed only 0.6% of children with covid-19 were critically ill [5].

A severe disease phenotype has emerged in children which appears temporally associated with SARS-CoV-2 infection [8,9]. The condition was first described in May 2020 in a cluster of children admitted to critical care in South London (UK) with evidence of a multisystem hyperinflammatory state with features similar to Kawasaki disease and toxic shock syndrome [8]. These children required inotropic support for refractory circulatory shock and mechanical ventilation for cardiovascular stabilisation rather than respiratory failure. Similar cohorts have been reported in Italy [10] and France [11]. The European Centre for Disease Prevention and Control on 15^th^ May 2020 estimated around 230 children had presented with this new syndrome in EU/EEA countries with two fatalities [3]. Preliminary case definitions have been proposed by the WHO [9] and the Royal College of Paediatrics and Child Health (RCPCH) [12]. The WHO use the term multisystem inflammatory syndrome in children and adolescents temporarily related to covid-19 (MIS-C) while RCPCH describe this illness as Paediatric Inflammatory Multisystem Syndrome temporally associated with SARS-COV-2 (PIMS-TS).

We aimed to characterise the features of children and young people (aged less than 19 years old) admitted to hospital in the UK with laboratory-confirmed SARS-CoV-2 from the International Severe Acute Respiratory and emerging Infection Consortium (ISARIC) WHO Clinical Characterisation Protocol UK (CCP-UK) cohort. Since our study enrolled patients prospectively from the beginning of the pandemic, we also had a unique opportunity to examine the emergence, timing, risk factors, clinical presentation, progression, course and outcomes of children and young people meeting the WHO preliminary case definition for MIS-C [9].

## Methods

### Study design and setting

The ISARIC WHO CCP-UK is an ongoing prospective cohort study across acute care hospitals in England, Wales and Scotland (8). This standing protocol for studying disease caused by pathogens of public health interest was activated on 17^th^ January 2020. The protocol, associated documents, and detail of the Independent Data and Material Access Committee (IDAMAC) are available at https://isaric4c.net. STROBE guidelines were used when reporting.

### Participants

Patients of any age admitted to hospital with proven or high likelihood of SARS-CoV-2 infection were enrolled into the ISARIC WHO CCP-UK cohort as previously described [13]. We present the data from children and young people aged less than 19 years old on the date of hospital admission, enrolled into the study up to and including 5^th^ June 2020, who had at least two weeks of outcome data available, and who had documented laboratory evidence of a positive result for SARS-CoV-2 (by PCR or serology). Patients who were already admitted for other clinical reasons and subsequently tested positive for SARS-CoV-2 whilst an inpatient were also included in this cohort.

### Data collection

Demographic and baseline data (including comorbidities and regular medications taken) alongside data pertaining to symptoms, clinical signs (including vital signs) during admission, laboratory and pathology investigations, treatments received whilst admitted and outcome were collected onto case report forms (supplement available online). Data regarding illness progression and severity, including location within the hospital (ward vs. critical care) were collected on day 1 (admission/diagnosis), days 3, 6, and 9, admission to critical care and on discharge/death. Data were collected from healthcare records onto the case report forms through a secure online database, REDCap (Research Electronic Data Capture, Vanderbilt University, hosted by the University of Oxford (UK)). Collection of this routine anonymised demographic and clinical data from medical records did not require consent in England and Wales. In Scotland, a waiver for consent was obtained from the Public Benefit and Privacy Panel.

### Variables

The case report form was agnostic to patient age and as such existing comorbidity variables were not tailored to the paediatric population (see **Supp Methods** for case report form and recoding of paediatric variables). Ethnicity was self-reported and transcribed from the healthcare record. The Paediatric Early Warning Score (PEWS) was used as a measure of disease severity at admission [14].

### Criteria for diagnosis of Multisystem Inflammatory Syndrome in Children and Adolescents (MIS-C)

The WHO preliminary case definition for MIS-C [9] was used as a framework for identifying children with the syndrome within this dataset (box 1).

#### Box 1.

WHO preliminary case definition for MIS-C with adaptations in italics to allow case identification in the ISARIC WHO CCP-UK cohort.

1. Fever for 3 days or more (*of any duration, self-reported prior to presentation*)
2. Plus <u>two</u> of the following:
  a. Rash or bilateral non-purulent conjunctivitis or muco-cutaneous inflammatory signs (*self-reported rash / conjunctivitis*)
  b. Hypotension or shock (*age < 2 years systolic blood pressure < 60 mmHg;* ≥*2 and < 5 years <70 mmHg;* ≥ *5 and < 12 years <80 mmHg;* ≥ *12 years < 90 mmHg at any point in admission* [14])
  c. Features of myocardial dysfunction, pericarditis, valvulitis, or coronary abnormalities (*diagnosis of endocarditis or myocarditis or documented pericardial effusion, coronary artery aneurysm, cardiomegaly or cardiac dysfunction on echocardiography*)
  d. Evidence of coagulopathy (*INR > 1*.*2 (any age* [15]*); age* ≤*3 months and PT >14s; age > 3 months and PT > 12s* [16])
  e. Acute gastrointestinal problems (*self-reported diarrhoea, vomiting, or abdominal pain*)
3. Plus elevated markers of inflammation (*CRP* ≥ *60 mg/L or ferritin* ≥ *200 mg/L at any point during admission; cut offs chosen after expert discussion)*
4. Plus no other obvious microbial cause of inflammation (*no significant positive growth on blood culture / cerebrospinal fluid culture during admission*)

### Outcomes

The primary outcomes of this study were critical care admission (high dependency unit (HDU) or intensive care unit (ICU)), development of MIS-C, and in-hospital mortality. Requirement for respiratory and cardiovascular support were also examined. A minimum two-week follow-up time was ensured for all included patients.

### Bias

Specialist children’s hospitals (tertiary-care) with paediatric-specific research teams may be over-represented. Capacity to enrol was also limited by staff resources at times of high covid-19 activity, and we were unable to comment on patients not recruited.

### Missing data

The pandemic disrupted routine care and usual research activities limiting data collection and verification, particularly during the peak of outbreak activity. We did not impute missing data for this descriptive analysis. To reduce the impact of missing data on outcome analyses, we restricted these analyses to patients who had been admitted for at least two weeks before data extraction. Complete data were not available for all variables, hence denominators differ between analyses.

### Statistical analysis

Continuous variables are displayed as mean (standard deviation) or median (interquartile range) if non-normally distributed. Categorical variables are presented as a frequency (percentage) unless otherwise stated. For univariable comparisons, we used Welch’s t, ANOVA, Mann–Whitney U, or Kruskal–Wallis tests according to data distribution. Categorical data were compared using chi-squared tests. A p-value of <0.05 was considered statistically significant; all tests were two sided. No adjustment was made for multiple comparisons. Parsimonious criterion-based model building used the following principles: relevant explanatory variables were identified from previous studies; interactions were checked at first order level; final model selection was informed by the Akaike Information Criterion (AIC) and c-statistic, with appropriate assumptions checked including the distribution of residuals. Analysis of symptom co-occurrence was performed using the Jaccard similarity coefficient and presented as a hierarchically ordered heatmap. Statistical analyses were performed using R (R Core Team version 3.6.3, Vienna, Austria) with packages including tidyverse, finalfit lubridate, ggplot2, gplot, dendextend and UpSetR.

### Patient and public involvement

This was an urgent public health research study in response to a Public Health Emergency of International Concern. Patients or the public were not involved in the design, conduct, or reporting of this rapid response research.

## Results

451 patients under 19 years with laboratory-confirmed SARS-CoV-2 (451/55 092, 0.8% of total cohort) were enrolled at 116 hospitals in England, Scotland, and Wales between 17 January and 5 June 2020, and 55 092 people of all ages were admitted to 260 hospitals. (Error! Reference source not found. **and Supp Figures 1 and 2**).

### Age, sex and ethnicity

The median age of the children was 3.9 years [IQR 0.3-12.9 years] and 57% (256/450) were male. Ethnicity was recorded in 89% (401/451) of cases: 56% (224/401) were White, 12% (49/401) were South Asian and 10% (40/401) were of Black ethnicity. At least one comorbidity was reported in 43% (195/451).

### Symptoms

The most common presenting symptoms were fever (73%, 306/418), cough (43%, 175/408), shortness of breath (32%, 124/389) and nausea/vomiting (32%, 120/380, Error! Reference source not found., **upper panel**). Fever, cough and shortness of breath did not show an association with age, however nausea and vomiting, abdominal pain, headache, and sore throat showed an increasing trend with age (**Supp Figure 4**). A heatmap and dendrogram of presenting symptoms revealed distinct clusters of clinical sub-phenotypes (Error! Reference source not found., **lower panel**). These comprised a respiratory illness, with clustering of both upper and lower respiratory symptoms together (light green and brown clusters) in addition to a cluster representing a muco-enteric illness (orange clusters). The latter cluster contained the symptoms specified in the WHO preliminary case definition for MIS-C (diarrhoea, abdominal pain, vomiting, rash and conjunctivitis) in addition to fatigue and headache. The two main clusters were not entirely dichotomous, and the muco-enteric cluster showed some overlap with upper respiratory tract symptoms (light green cluster), but very little overlap with the lower respiratory tract symptoms of wheeze and lower chest wall indrawing (brown cluster).

### Comorbidities

The most common comorbidities were neurological (12%, 51/419), haematological/oncological/immunological (combined category as described in **Supp Methods**, 8%, 35/420) and cardiac (7%, 31/416). Data on prematurity (defined as being born before completion of 37 weeks gestation) was only routinely collected for children aged under 1 year old and 22% (32/148) were premature (**Supp Table 1 and Supp Figure 3**).

The median PEWS at presentation was 3 [IQR 1.0-5.0] and blood tests at presentation were mainly within normal ranges (**Supp Tables 2 and 3**). Antibiotics were given to 71% (281/397) of children and 7% (26/368) received antiviral medication (22 received acyclovir and 4 received remdesivir, **Table 2**).

**Table 1.** Demographics across the cohort of patients under 19 years with laboratory confirmation of SARS-CoV-2. Immunosuppressant use includes oral but not inhaled corticosteroids. Prematurity defined as birth before completion of 37 weeks gestation.

| Variable | Level | n (%) |
| --- | --- | --- |
| Total N (%) |  | 451 (100.0) |
| Age on admission (years) | Median (IQR) | 3.9 (0.3 to 12.9) |
| Age (years) | < 1 m | 30 (6.7) |
|  | ≥1 m <1 yr | 132 (29.3) |
|  | 1-4 | 74 (16.4) |
|  | 5-9 | 65 (14.4) |
|  | 10-14 | 67 (14.9) |
|  | 15-19 | 83 (18.4) |
| Sex at Birth | Male | 256 (56.8) |
|  | Female | 194 (43.0) |
|  | (Missing) | 1 (0.2) |
| Ethnicity | White | 224 (49.7) |
|  | Black | 40 (8.9) |
|  | East Asian | 10 (2.2) |
|  | South Asian | 49 (10.9) |
|  | Other | 78 (17.3) |
|  | (Missing) | 50 (11.1) |
| Admitted > 5 days before symptom onset | No | 385 (85.4) |
|  | Yes | 42 (9.3) |
|  | (Missing) | 24 (5.3) |
| Any comorbidity | No/Unknown | 256 (56.8) |
|  | Yes | 195 (43.2) |
| Neurological | No | 368 (81.6) |
|  | Yes | 51 (11.3) |
|  | (Missing) | 32 (7.1) |
| Haematology / Oncology / Immunology | No | 385 (85.4) |
|  | Yes | 35 (7.8) |
|  | (Missing) | 31 (6.9) |
| Prematurity | No | 116 (25.7) |
|  | Yes | 32 (7.1) |
|  | (Missing) | 303 (67.2) |
| Immunosuppressant use prior to presentation | No | 372 (82.5) |
|  | Yes | 38 (8.4) |
|  | (Missing) | 41 (9.1) |

**Table 2.**
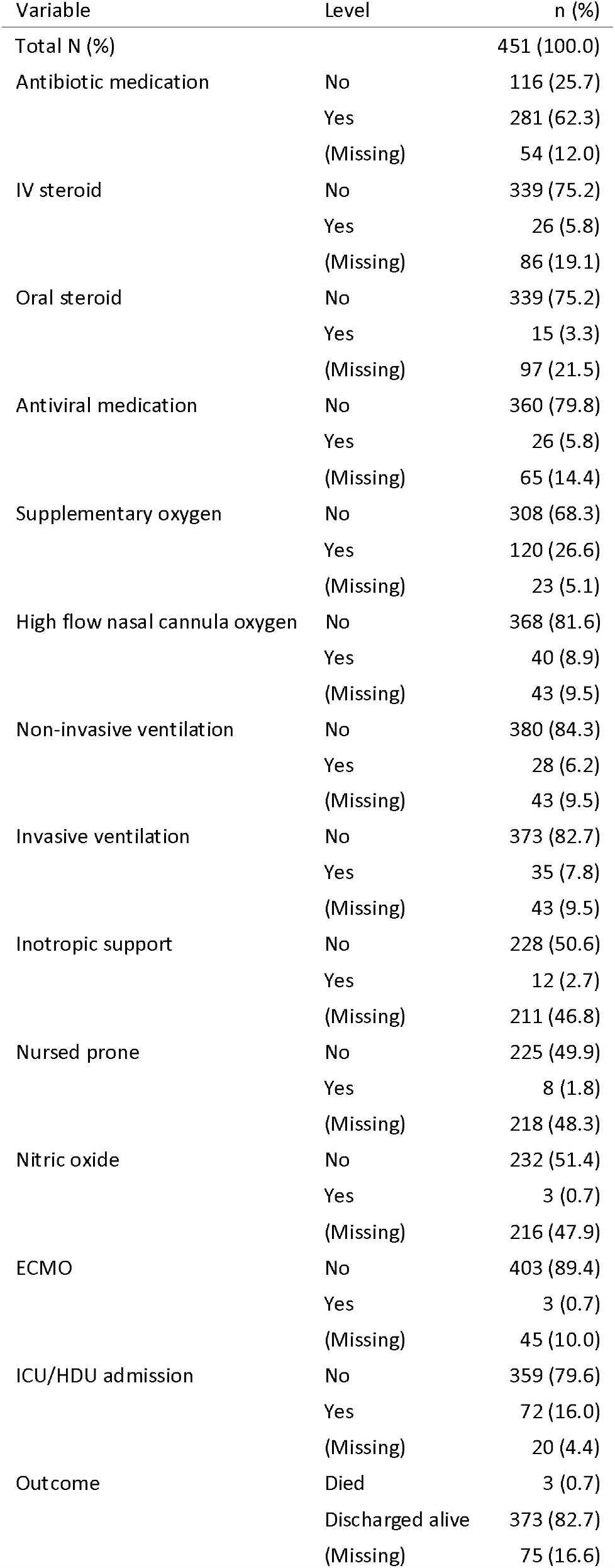
Treatments received and outcomes. ICU/HDU = Intensive care unit / high dependency unit. ECMO = Extracorporeal membrane oxygenation

| Variable | Level | n (%) |
| --- | --- | --- |
| Total N (%) |  | 451 (100.0) |
| Antibiotic medication | No | 116 (25.7) |
|  | Yes | 281 (62.3) |
|  | (Missing) | 54 (12.0) |
| IV steroid | No | 339 (75.2) |
|  | Yes | 26 (5.8) |
|  | (Missing) | 86 (19.1) |
| Oral steroid | No | 339 (75.2) |
|  | Yes | 15 (3.3) |
|  | (Missing) | 97 (21.5) |
| Antiviral medication | No | 360 (79.8) |
|  | Yes | 26 (5.8) |
|  | (Missing) | 65 (14.4) |
| Supplementary oxygen | No | 308 (68.3) |
|  | Yes | 120 (26.6) |
|  | (Missing) | 23 (5.1) |
| High flow nasal cannula oxygen | No | 368 (81.6) |
|  | Yes | 40 (8.9) |
|  | (Missing) | 43 (9.5) |
| Non-invasive ventilation | No | 380 (84.3) |
|  | Yes | 28 (6.2) |
|  | (Missing) | 43 (9.5) |
| Invasive ventilation | No | 373 (82.7) |
|  | Yes | 35 (7.8) |
|  | (Missing) | 43 (9.5) |
| Inotropic support | No | 228 (50.6) |
|  | Yes | 12 (2.7) |
|  | (Missing) | 211 (46.8) |
| Nursed prone | No | 225 (49.9) |
|  | Yes | 8 (1.8) |
|  | (Missing) | 218 (48.3) |
| Nitric oxide | No | 232 (51.4) |
|  | Yes | 3 (0.7) |
|  | (Missing) | 216 (47.9) |
| ECMO | No | 403 (89.4) |
|  | Yes | 3 (0.7) |
|  | (Missing) | 45 (10.0) |
| ICU/HDU admission | No | 359 (79.6) |
|  | Yes | 72 (16.0) |
|  | (Missing) | 20 (4.4) |
| Outcome | Died | 3 (0.7) |
|  | Discharged alive | 373 (82.7) |
|  | (Missing) | 75 (16.6) |

### Children requiring critical care

17% of children (72/431) were admitted to critical care (ICU or HDU level care); 5% (12/240) received inotropic support, 7% (28/408) received non-invasive ventilation and 9% (35/408) received invasive mechanical ventilation (**Tables 2, Table 3 and Supp Figure 5**). Black ethnicity was significantly associated with admission to critical care on multivariable analysis (odds ratio (OR) 3.02 (95% confidence interval 1.30 to 6.84, p=0.008 **Table 4 and Supp Table 4**). On multivariable analysis, both age under one month (OR 5.05 (1.69 to 15.72, P=0.004)) and age between 10-14 years (OR 3.11 (1.21 to 8.55, p=0.022)) were associated with admission to critical care (reference age group 15 to 19 years), but there was no association with sex on either univariable or multivariable analysis (**Table 3, Table 4 and Supp Table 4**).

**Table 3.**
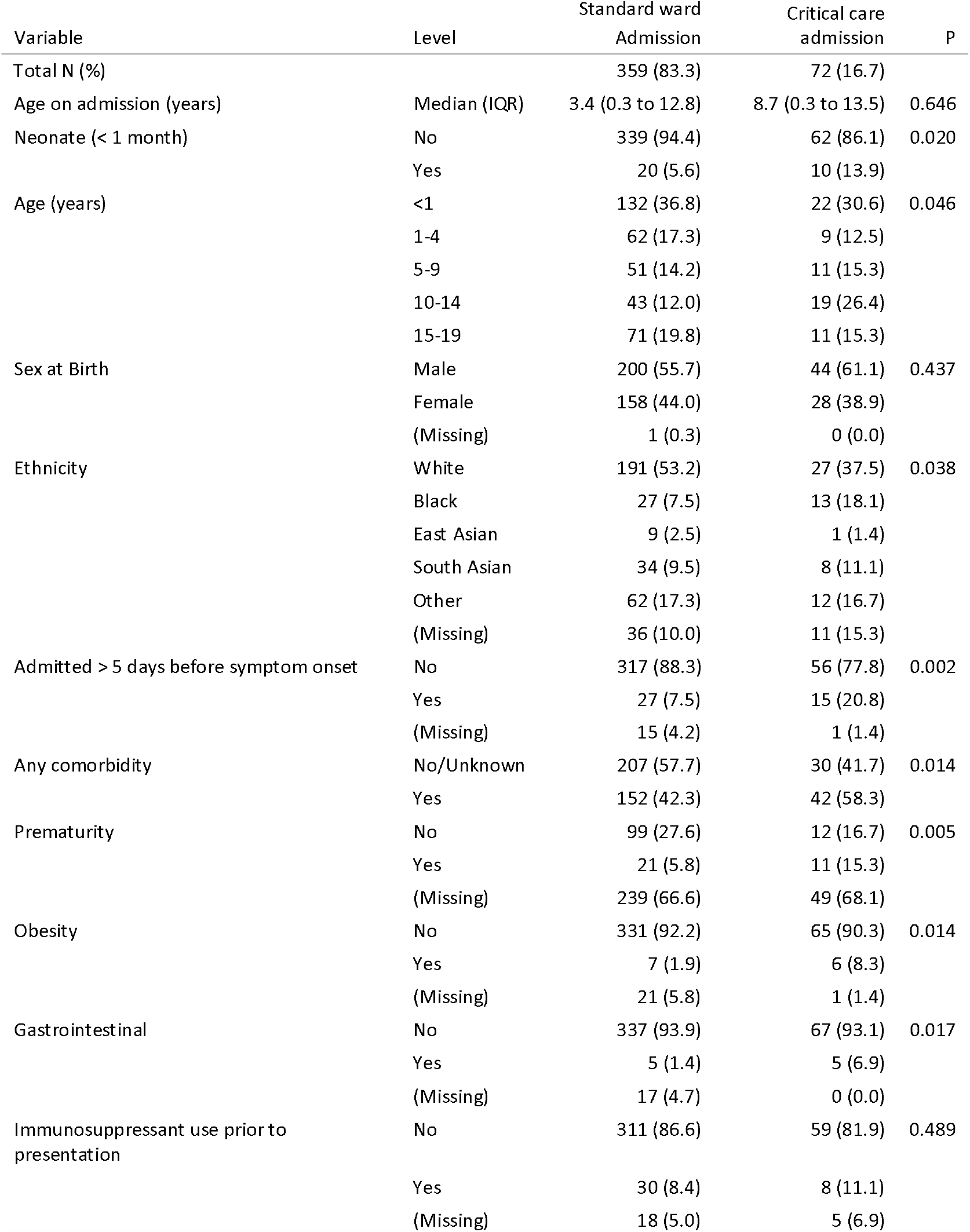
Demographics stratified by admission to critical care. Categorical variables analysed using Fisher’s exact test, continuous variables by Kruskal Wallis test.

| Variable | Level | Standard ward<br>Admission | Critical care<br>admission | P |
| --- | --- | --- | --- | --- |
| Total N (%) |  | 359 (83.3) | 72 (16.7) |  |
| Age on admission (years) | Median (IQR) | 3.4 (0.3 to 12.8) | 8.7 (0.3 to 13.5) | 0.646 |
| Neonate (< 1 month) | No | 339 (94.4) | 62 (86.1) | 0.020 |
|  | Yes | 20 (5.6) | 10 (13.9) |  |
| Age (years) | <1 | 132 (36.8) | 22 (30.6) | 0.046 |
|  | 1-4 | 62 (17.3) | 9 (12.5) |  |
|  | 5-9 | 51 (14.2) | 11 (15.3) |  |
|  | 10-14 | 43 (12.0) | 19 (26.4) |  |
|  | 15-19 | 71 (19.8) | 11 (15.3) |  |
| Sex at Birth | Male | 200 (55.7) | 44 (61.1) | 0.437 |
|  | Female | 158 (44.0) | 28 (38.9) |  |
|  | (Missing) | 1 (0.3) | 0 (0.0) |  |
| Ethnicity | White | 191 (53.2) | 27 (37.5) | 0.038 |
|  | Black | 27 (7.5) | 13 (18.1) |  |
|  | East Asian | 9 (2.5) | 1 (1.4) |  |
|  | South Asian | 34 (9.5) | 8 (11.1) |  |
|  | Other | 62 (17.3) | 12 (16.7) |  |
|  | (Missing) | 36 (10.0) | 11 (15.3) |  |
| Admitted > 5 days before symptom onset | No | 317 (88.3) | 56 (77.8) | 0.002 |
|  | Yes | 27 (7.5) | 15 (20.8) |  |
|  | (Missing) | 15 (4.2) | 1 (1.4) |  |
| Any comorbidity | No/Unknown | 207 (57.7) | 30 (41.7) | 0.014 |
|  | Yes | 152 (42.3) | 42 (58.3) |  |
| Prematurity | No | 99 (27.6) | 12 (16.7) | 0.005 |
|  | Yes | 21 (5.8) | 11 (15.3) |  |
|  | (Missing) | 239 (66.6) | 49 (68.1) |  |
| Obesity | No | 331 (92.2) | 65 (90.3) | 0.014 |
|  | Yes | 7 (1.9) | 6 (8.3) |  |
|  | (Missing) | 21 (5.8) | 1 (1.4) |  |
| Gastrointestinal | No | 337 (93.9) | 67 (93.1) | 0.017 |
|  | Yes | 5 (1.4) | 5 (6.9) |  |
|  | (Missing) | 17 (4.7) | 0 (0.0) |  |
| Immunosuppressant use prior to presentation | No | 311 (86.6) | 59 (81.9) | 0.489 |
|  | Yes | 30 (8.4) | 8 (11.1) |  |
|  | (Missing) | 18 (5.0) | 5 (6.9) |  |

**Table 4.**
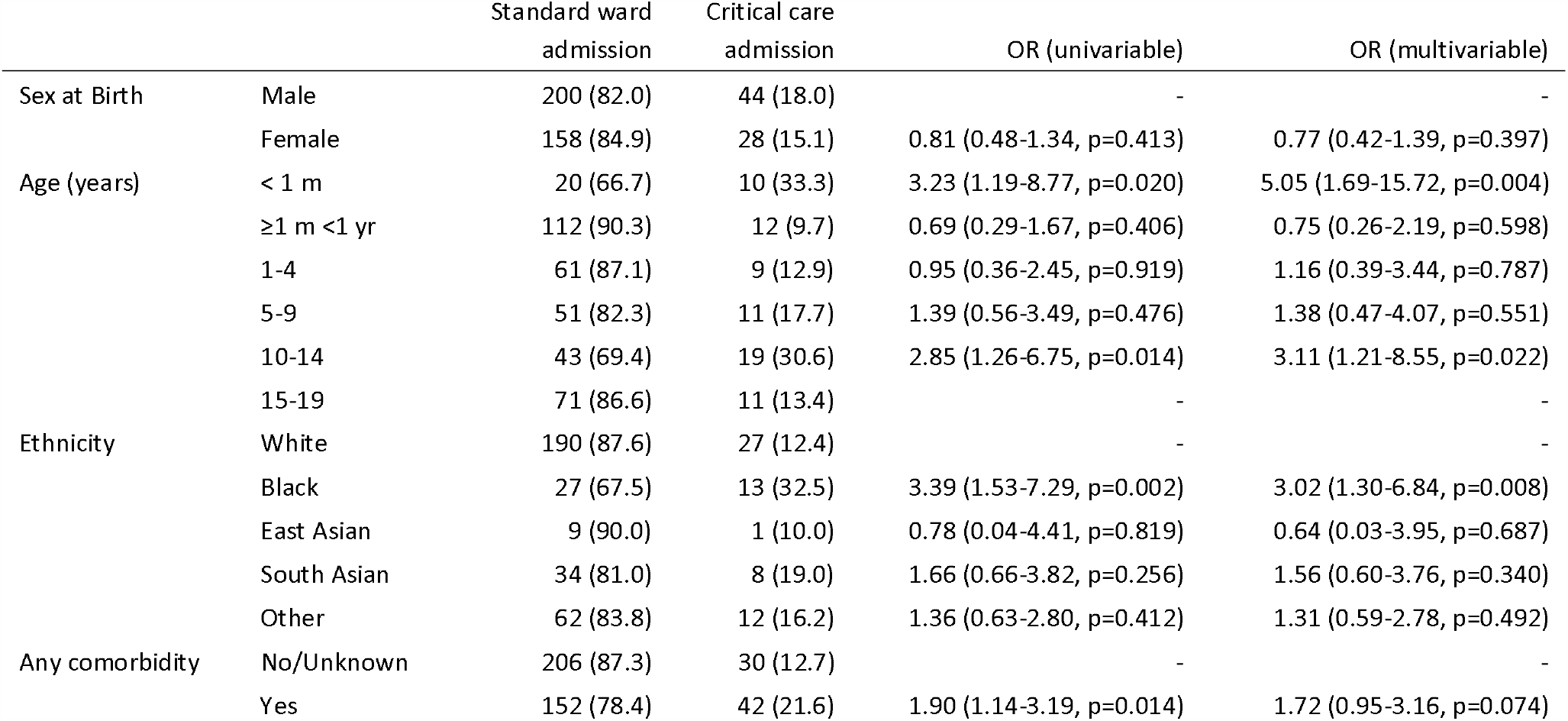
Factors associated with admission to critical care unit. Univariable and multivariable logistic regression analyses were performed using potential predictors identified a priori and during exploratory analyses. Results are odds ratios with 95% confidence intervals. Data are otherwise n (%). OR, odds ratio; m, month; yr, year;. OR, odds ratio with 95% confidence intervals

On univariable analysis, children with comorbidities were more likely to be admitted to critical care than those without (OR 1.90 (1.14 to 3.19, p=0.014) however, this no longer reached significance in the multivariable model (OR 1.72 (0.95 to 3.26, p = 0.074). Comorbidities most commonly associated with critical care admission on univariable analysis were prematurity (48% (11/23) critical care admissions vs 18% (21/120) of standard care admissions, p=0.005), obesity (8% (6/71) vs 2% (7/338), p=0.014) and gastrointestinal comorbidities (7% (5/72) vs 1% (5/342), p=0.017, **Table 3 and Supp Table 5**). Children receiving critical care were more likely to have been admitted to hospital more than 5 days before their symptoms started (indicating likely hospital acquired infection) than those receiving ward level care (15/71 (21%) vs 27/344 (8%) p=0.002). No association was seen between prior immunosuppressant use and critical care admission (Error! Reference source not found.).

Children admitted to critical care were more likely to have presented with diarrhoea, (35% (22/63) vs 12% (36/309), p <0.001), conjunctivitis (13% (8/62) vs 2% (6/285), p=0.001) and confusion (22% (13/60) vs 7% (20/302), p=0.001) than those cared for on a standard ward (**Supp Table 6**). They were objectively more unwell at presentation than those receiving standard ward care (median PEWS of 5 [IQR 3.0-8.0] vs 2 [1.0-4.0], p<0.001, **Supp Table 7**). There were also significant differences in haematologic, biochemical and radiologic abnormalities between the two groups at presentation (**Supp Table 8**). In particular, children admitted to critical care had a lower platelet count (median 206.5 ×10^9^/L [IQR 129.0-296.8] vs 298.0 ×10^9^/L [233.0-387.0], p <0.001), a higher neutrophil count (7.5 ×10^9^/L [4.8-13.0] vs 4.6 ×10^9^/L [2.3-8.3], p <0.001) and higher CRP (66.0mg/L [7.0-211.3] vs 12.0mg/L [4.0-58.0], p<0.001) at presentation than those cared for on a standard ward. Children admitted to critical care were also more likely to have infiltrates on chest radiograph (64% (32/50) vs 34% (38/112), p <0.001).

### Patient outcomes

Overall, 83% (373/451) of children and young people were discharged alive, 0.7% (3/451) died, and 17% (75/451) continued to receive care at the date of reporting or were missing outcomes (**Table 2**).

There were no deaths in children under 16 years of age in this cohort. Three young people died, who were 16 to 19 years. Two of these young people had profound neurodisability with pre-existing respiratory compromise. The third young person was immunosuppressed by chemotherapy for a haematological malignancy.

### Patients meeting the WHO preliminary case definition for MIS-C

Twelve per cent (36/303) of children met the WHO preliminary case definition for MIS-C (17) (**Table 5 and Supp Figure 6**). The first patient identified developed symptoms in mid-March, when covid-19 cases were increasing nationally, followed by a small number of cases identified steadily throughout the surveillance period (**Figure 2**). Geographically, the highest number of children with MIS-C came from areas with the largest covid-19 outbreaks, namely the Midlands and London (**Supp Figure 7**).

**Table 5.**
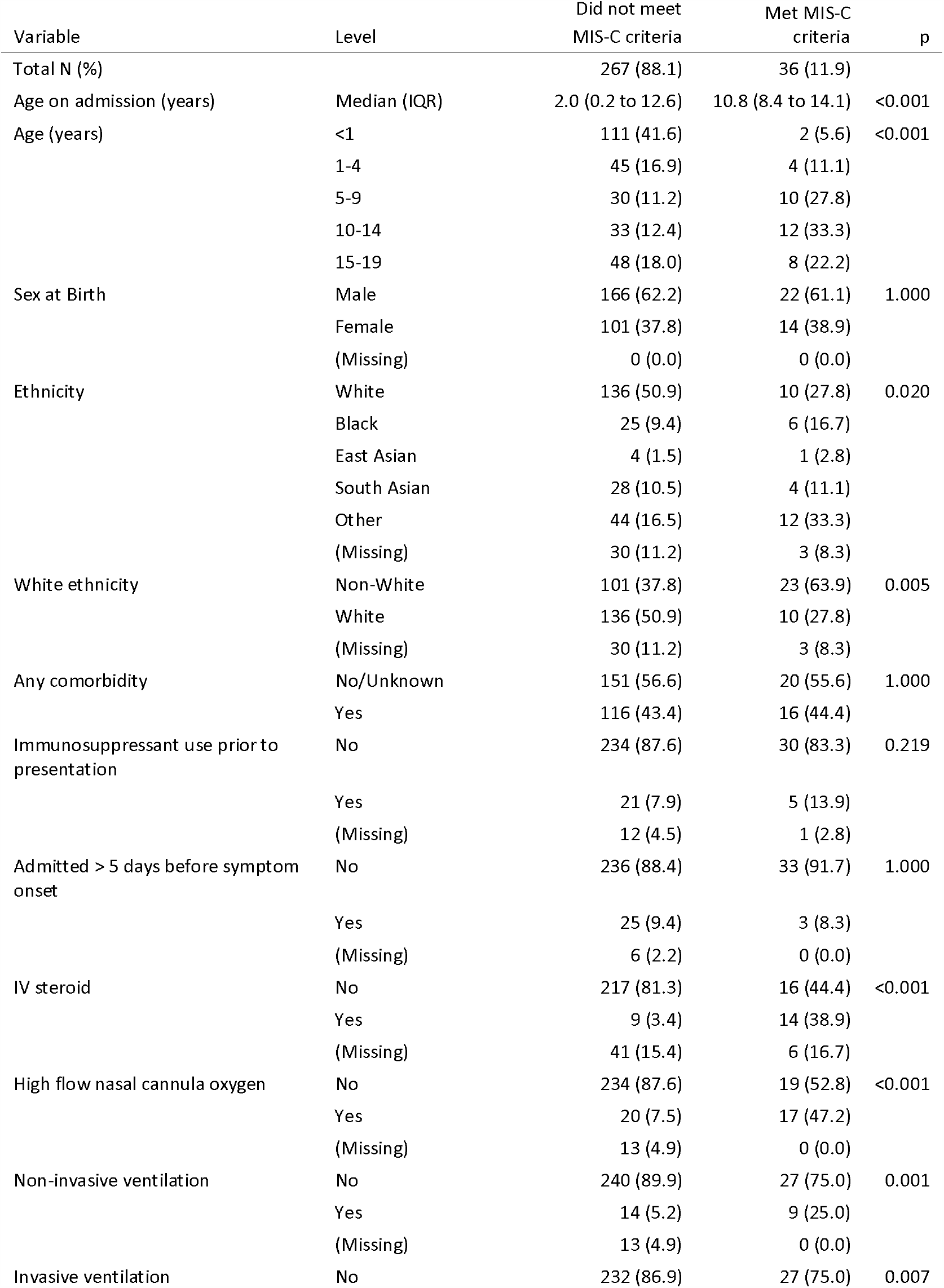
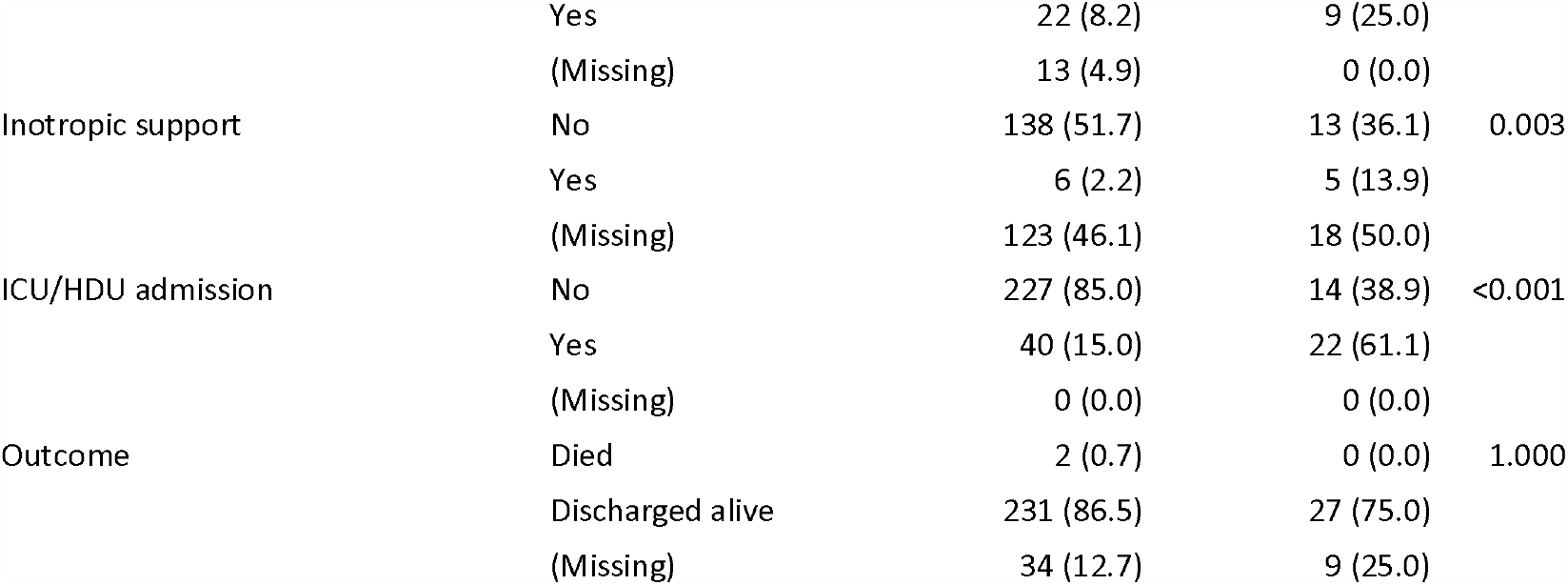
Demographics at presentation and therapies administered stratified by Multisystem Inflammatory Syndrome in Children and Adolescents (MIS-C) status. Categorical variables analysed using Fisher’s exact test, continuous variables by Kruskal Wallis test.

**Figure 1.**
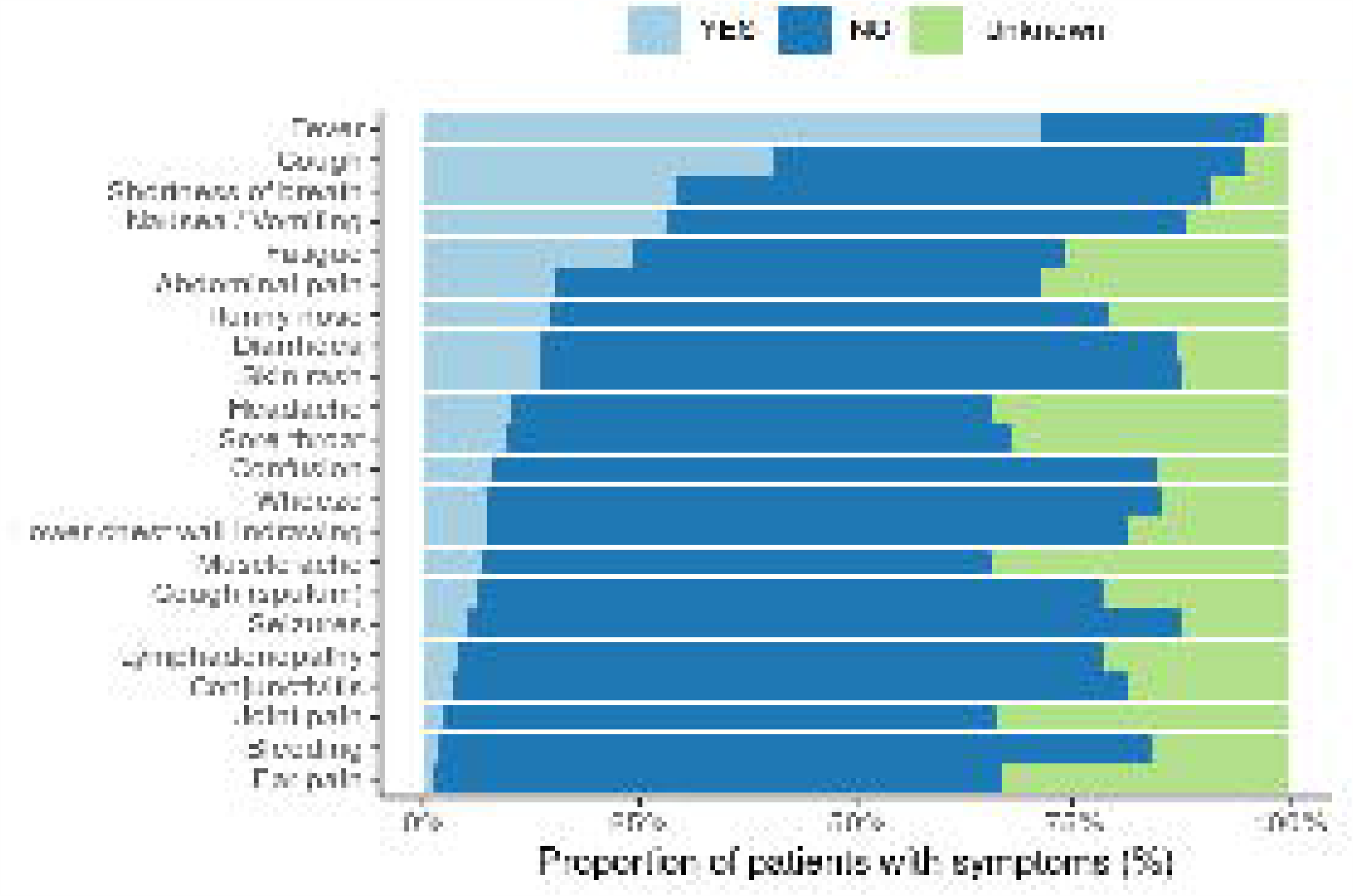
Upper Panel. Proportion of patients presenting with each symptom.

**Figure 1a.**
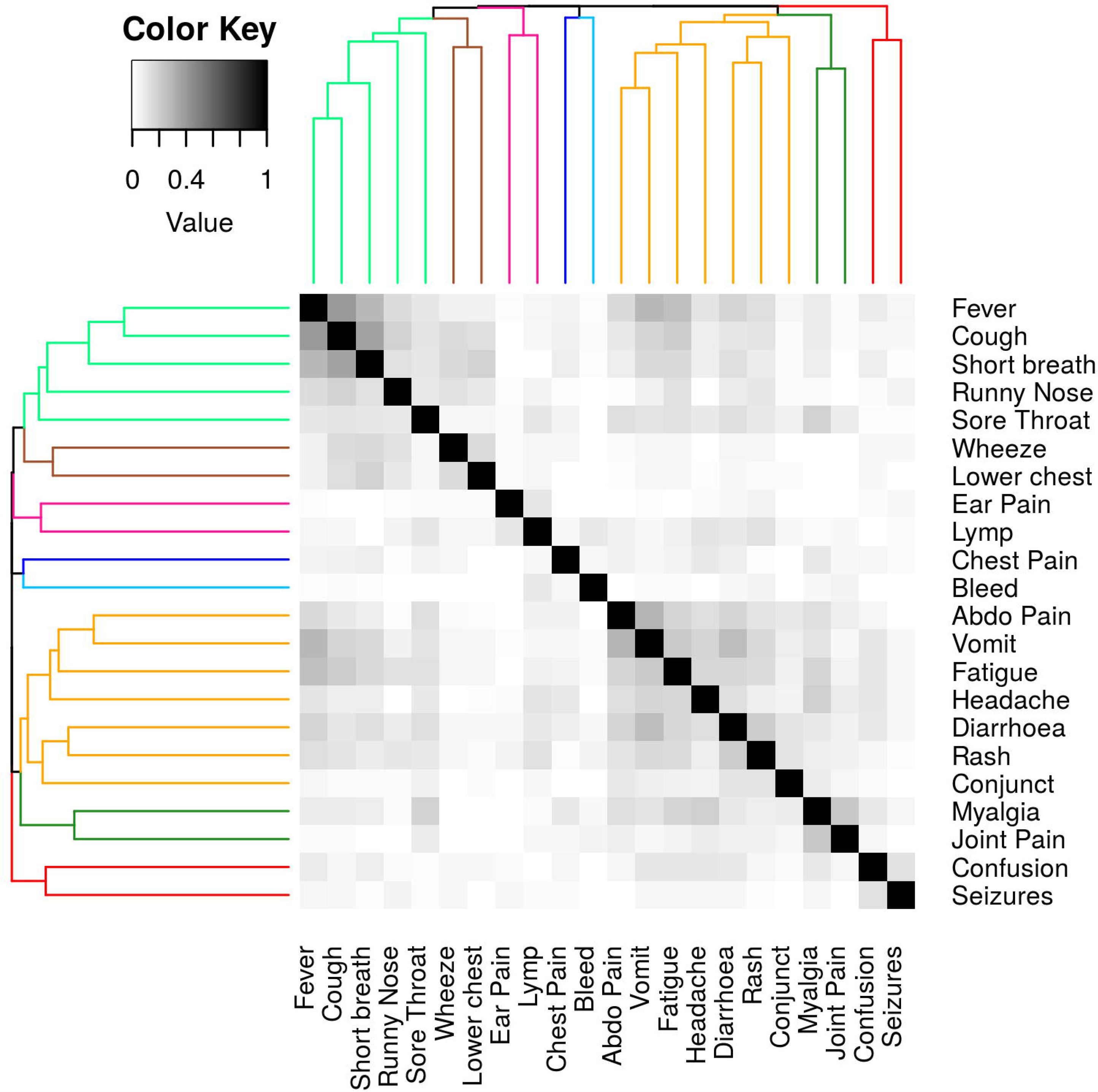
Lower Panel. Heatmap with dendrogram describing the clusters of co-occurring symptoms calculated using hierarchical clustering with Jaccard distance as metric and complete linkage. Heatmap displays the pairwise Jaccard indices among 22 symptoms. Jaccard index is a measure of similarity that computes the ratio of the number of times two symptoms appear together in the data and the number of times either of them appear in the data. The index varies between 0 and 1 with 0 implying that the two symptoms never appear together (no co-occurrence), and 1 implying that the two symptoms only appear together (co-occurrence only). The dendrogram describing the clusters of symptoms in the heatmap was calculated using hierarchical clustering with Jaccard distance as metric and complete linkage, where Jaccard distance is calculated by subtracting the Jaccard index from 1. Conjunc = conjunctivitis. Lower chest = lower chest wall indrawing. Lymp = lymphadenopathy

**Figure 2.**
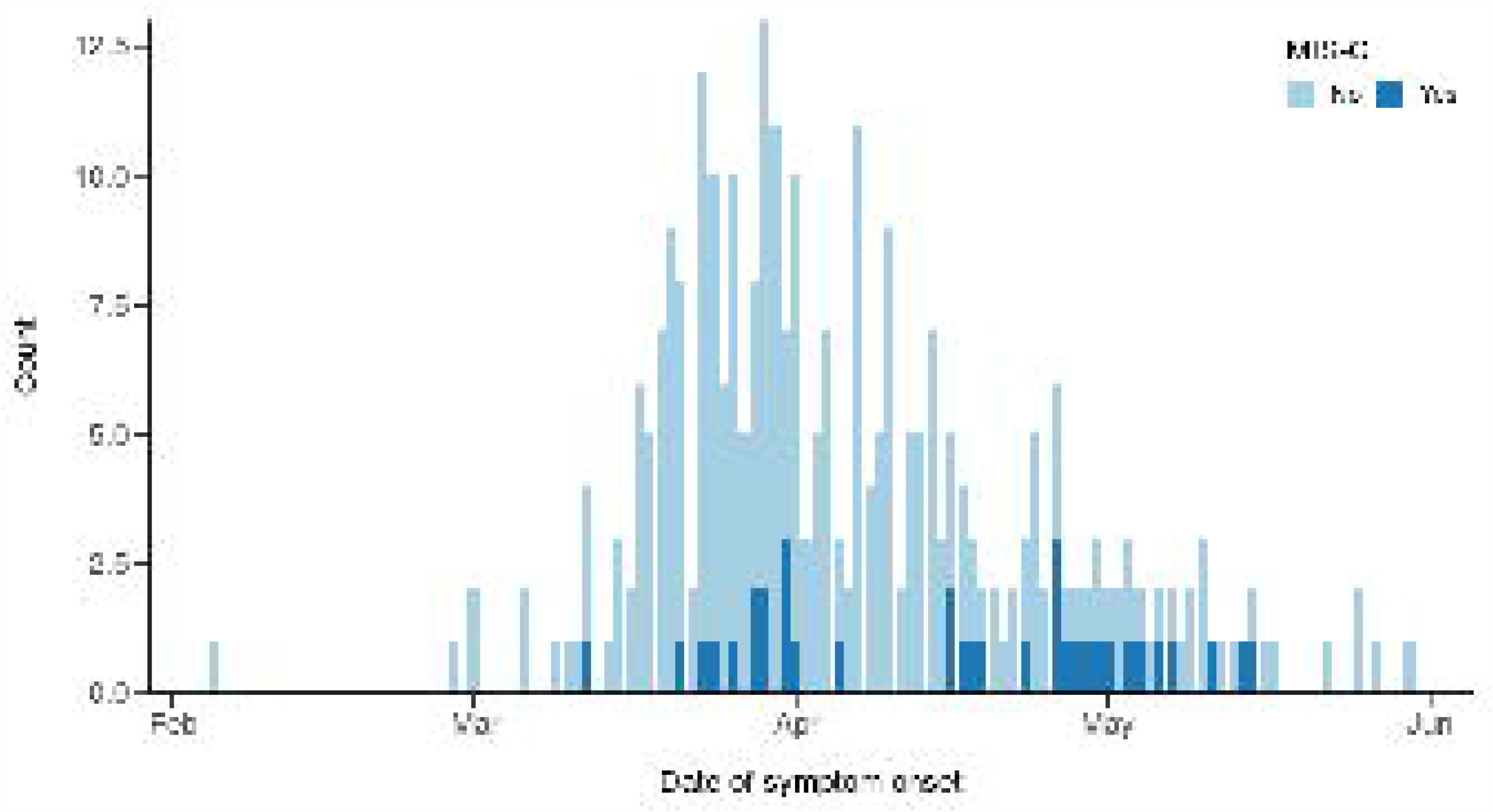
Dates of symptom onset for paediatric cases of SARS-CoV-2 infection and cases meeting the WHO preliminary criteria for Multisystem Inflammatory Syndrome in Children and Adolescents (MIS-C) in the ISARIC WHO CCP-UK cohort over time.

Children meeting the WHO preliminary definition for MIS-C were significantly older than those who did not (median 10.8 years [IQR 8.4-14.1] vs 2.0 [0.2-12.6], p<0.001) and were significantly more likely to be of non-White ethnicity (70% (23/33) vs 43% (101/237), p=0.005, **Table 5**). MIS-C was not associated with comorbidity (**Supp Table 9**). In addition to the WHO case definition features (fever, rash, conjunctivitis and gastrointestinal symptoms), the children with MIS-C were also more likely to present with headache (45% (13/29) vs 11% (19/171), p<0.001), myalgia (39% (11/28) vs 7% (12/170), p<0.001) lymphadenopathy (21% (6/29) vs 3% (6/210), p=0.001), sore throat (37% (10/27) vs (13% (24/183), p = 0.004) and fatigue (57% (17/30) vs 31% (60/192), p=0.012) than children who did not (**Supp Table 10**). Children with MIS-C also had a higher PEWS score at presentation (median 5.5 [IQR 3.0-7.0] vs 3.0 [1.0-5.0], p<0.001) and were more likely to have reduced consciousness (32% (10/31) vs 9% (17/200), p=0.001) than those who did not meet the criteria (**Supp Table 11 and Supp Figure 8**).

Review of laboratory investigations found that children meeting the criteria for MIS-C were more likely to have a platelet count of less than 150 ×10^9^/L than those who did not (30% (10/33) vs 10% (24/232), p=0.004, **Supp Table 12**). Children meeting the MIS-C criteria also had lower lymphocyte counts (median 0.9 × 10^9^/L [IQR 0.7-1.8] vs 2.1 × 10^9^/L [1.4-3.6], p<0.001) but higher neutrophil counts (7.8 × 10^9^/L [5.7-12.3] vs 5.1 × 10^9^/L [2.3-9.3], p=0.001) and higher creatinine (57.0 μmol/L [38.0-74.8] vs 28.0 μmol/L [20.0-46.0], p<0.001) than those without (**Supp Table 12**).

Children with MIS-C were four times more likely to be admitted to critical care (61% (22/36) vs 15% (40/267, p<0.001, **Table 5**). They were more likely to receive intravenous corticosteroids (47%, (14/30) vs 4% (9/226) p <0.001), non-invasive (25% (9/36) vs 6% (14/254), p=0.001) and invasive ventilation (25% (9/36) vs 9% (22/254), p=0.007) as well as inotropic support (28% (5/18) vs 4% (6/144), p=0.003, **Table 5**). There were no deaths in the MIS-C group.

## Discussion

### Principal Findings

There were 451 children and young people under 19 years with laboratory confirmed SARS-CoV-2 recruited to the ISARIC WHO CCP-UK study between 17 January and 5 June 2020, accounting for 0.8% of all patients in the whole cohort at that time. The median age of children with covid-19 was 3.9 years [IQR 0.3-12.9]. The cohort was predominantly male (57%) and of White ethnicity (56%) with most (57%) children having no known comorbidities. The most common presenting symptoms were fever, cough, shortness of breath, nausea and vomiting and a clear muco-enteric cluster of symptoms was seen. Seventeen percent of hospitalised children required critical care. Critical care admission was associated with age younger than one month, age 10 to 14 years and Black ethnicity. The in-hospital case fatality rate was strikingly low (3 patients, 0.7%) when compared with 25% (14 025 / 55 092) across all age groups in the cohort with covid-19 for the same time period. In this paediatric cohort, 12% of patients met the WHO preliminary criteria for MIS-C, which was associated with older age, non-White ethnicity and admission to critical care. MIS-C cases were first identified in mid-March when cases of covid-19 began to rise in the UK. In addition to the clinical criteria provide by the WHO, we found children with MIS-C were more likely to present with headache, myalgia, fatigue, sore throat and lymphadenopathy as well as a lower platelet count than children with SARS-CoV-2 who did not meet the MIS-C criteria.

### Strengths and limitations of this study

This study is unique in that data for patients with laboratory-confirmed covid-19 were collected prospectively and throughout the admission. The ISARIC WHO CCP-UK study had previously been activated in 2016 and 2018 for cases of Middle East Respiratory Syndrome (MERS) and monkeypox so was prepared for the SARS-CoV-2 pandemic, allowing swift activation. Consequently, in addition to reporting the clinical characteristics, risk factors and outcomes of covid-19 in children, this dataset provided a unique opportunity to objectively monitor the emergence and progression of a novel multisystem inflammatory syndrome in the UK, whilst minimising recall bias. The first patient meeting the criteria for MIS-C was identified on 19 March 2020, whilst the first published cases were reported on 6 May 2020 [8]. Comparison with overall covid-19 cases confirms the sporadic occurrence of the MIS-C throughout the first peak of the covid-19 pandemic in the UK. In contrast to previous reports, our analysis was limited only to children admitted with laboratory confirmed SARS-CoV-2 which allowed us to clearly define the picture of covid-19 in children and reduce confounding with other potential causes.

The ISARIC WHO CCP-UK database is estimated to represent 53% of covid-19 hospitalisations across England, Wales and Scotland. It is therefore susceptible to selection bias, particularly as tertiary centres with critical care units and specialist children’s hospitals are more likely to have dedicated research teams, potentially skewing the severity and age of the patients reported.

The most common presenting symptoms in children in our study (fever, cough and dyspnoea) reflect the original case definition for SARS-CoV-2 testing in the UK, suggesting that this paediatric cohort is likely to have been influenced by the testing criteria.

The PEWS is validated up to 16 years of age [14]. As ranges of clinical observations do not vary considerably between 16 and 18 year olds [17] PEWS scoring was extended to all those under 19 years. To identify children and young people meeting the WHO criteria for MIS-C it is necessary to have data on CRP and fever. Decisions to measure CRP and other parameters were at physicians’ discretion. Children missing either of these variables were excluded from this analysis.

A limitation of this study is the use of a case record form that was agnostic to age so not specifically tailored for paediatric data collection, particularly in regard to comorbidities. Some of this information was available in free text but these data were incomplete. By design we are not able to differentiate between people whose symptoms were directly attributable to SARS-CoV-2 infection and those who had been admitted for other reasons and then found to be positive for the virus. The study relied primarily on PCR testing as evidence of SARS-CoV-2 infection as diagnostic serology was not widely available at the start of the pandemic. As other studies of MIS-C report several children who are PCR-negative but have serological evidence of infection, this could have limited early recruitment of MIS-C cases. Finally, in order to share findings from this study promptly as an urgent public health research priority, these analyses were performed on a cohort with ongoing data collection and missing data, the proportion of which will decrease with time. We do not have data for children identified as infected with SARS-CoV-2 in the community who were not admitted to hospital, and we cannot yet report on sequelae of covid-19 in children after discharge.

### Comparison with other studies

Children and young people under 19 years accounted for 0.8% (451/55 394) of the ISARIC WHO CCP-UK cohort on 5 June 5 2020, which is broadly consistent with 2% reported in China [1] and 1.7% in North America [2]. Our cohort of paediatric hospitalised cases had a median age of 3.9 years which was similar to an Italian cohort (3.3 years [18]), but younger than Chinese (6-7 years) [5,6] and North American (11 years) cohorts [2], however these other cohorts were not limited to hospitalised children. While respiratory presentations were most common, 35% of children also had gastrointestinal symptoms at presentation, which is higher than 10-22% reported in other paediatric literature [2,18,19]. Gastrointestinal symptoms have also been prominent in children presenting with infection by MERS-CoV (28%) [20] and severe acute respiratory syndrome coronavirus (30%) [21]. We also identify for the first time a clear muco-enteric cluster of symptoms which shows some overlap with upper, but not lower respiratory tract symptoms.

Children of Black ethnicity were over-represented, comprising 10% of our paediatric cohort compared with 4.7% of all children under 18 years across England and Wales [22] and 1% in Scotland [23]. This finding may also be influenced by the ethnic composition of the population served by the sites recruiting to this study. Black ethnicity was also associated with increased odds of admission to critical care on multivariable analysis, consistent with reports for adult populations suggesting South Asian and Black ethnicities are disproportionately severely affected by SARS-CoV-2 infection [24–26]. Studies of paediatric covid-19 from other countries have either been from ethnically homogenous groups or have not reported ethnicity, making comparisons difficult.

The critical care admission rate in our cohort was 17% compared to 10% reported in a north American cohort of hospitalised children [2] and 13% in a multicentre cohort study across 25 European countries [19]. As previously noted, this rate may be elevated in our study owing to hospitals with dedicated paediatric research teams being more likely to provide paediatric critical care. The prevalence of comorbidities (53%) in children admitted to critical care in our cohort was also similar to that reported in the European multicentre study countries (52% [19]). Obesity was associated with critical care admission in our paediatric cohort, in agreement with adult data from ISARIC WHO CCP-UK [13]. In England 20% of children are obese by 11 years of age [27]. Childhood obesity, however, is also influenced by deprivation [27], which was not analysed in our study. Age under 1 month was associated with increased odds of critical care admission, in agreement with the European cohort [19]. As previously noted, a limitation of our study was the difficulty in differentiating between children with symptoms directly attributable to SARS-CoV-2 infection and those admitted for other reasons and subsequently testing positive for the virus. This may explain the association between age under one month and admission to critical care, if these babies were already admitted to neonatal intensive care and undergoing regular SARS-CoV-2 screening. Children who had been admitted to hospital for more than 5 days prior to symptoms were also more likely to be admitted to critical care. By definition, this group includes children with comorbidity. SARS-CoV-2 nosocomial infections in children are not well reported and this area requires closer scrutiny, ideally with the use of viral sequence data.

Using adapted WHO criteria [9], we identified 36 patients meeting the criteria for multisystem inflammatory syndrome. Initial UK reports described children admitted to hospital with circulatory shock and a hyperinflammatory state with features of similar to toxic shock or Kawasaki disease [8]. Children fulfilling the case definition for MIS-C have been reported in multiple regions experiencing large outbreaks of covid-19, including England (UK) [28], Paris (France) [11], Bergamo (Italy) [10] and New York City (USA) [29]. Ours is the first report, however, to identify cases and timelines using a prospective national data collection strategy. MIS-C appears to be temporally associated with covid-19 but a causal relationship remains to be established. Older age and non-White ethnicity were associated with MIS-C in our study, in agreement with a recent case series of 99 children with MIS-C from New York State (USA) where 63% were of non-White ethnicity and 69% were between 6 and 20 years [29]. Children in our study with MIS-C were much more unwell than other children with covid-19, with 28% requiring inotropic support, (compared with 20% in the Italian cohort[10], 47% in the French cohort [11] and 62% in the New York cohort [29].

Across the whole of our paediatric cohort, we identified a distinct cluster presenting with muco-enteric symptoms (rash, conjunctivitis, diarrhoea, vomiting and abdominal pain) in addition to headache and fatigue which overlapped very closely with the WHO preliminary case definition [9], suggesting that children with MIS-C may be at the severe end of this muco-enteric spectrum. The significant associations between MIS-C and headache, myalgia, fatigue, sore throat and lymphadenopathy in our cohort may be useful in refining the case definition. In addition, the association of MIS-C with platelet count less than 150 ×10^9^/L and low lymphocyte counts is in agreement with previous reports [28,29]. These important findings may assist in differentiating this syndrome from other illnesses, particularly Kawasaki disease where platelet counts are typically elevated.

### Conclusion and policy implications

Our data confirms less severe covid-19 in children and young people with SARS-CoV-2 infection than in adults. Admission to critical care was associated with age under 1 month, age 10-14 years and Black ethnicity. In agreement with previous reports, we find older age and non-White ethnicity to be associated with MIS-C. In addition, we provide evidence for refining the case definition for MIS-C including an association with low platelet count, headache, myalgia, lymphadenopathy, sore throat and fatigue which could be incorporated into future iterations of the case definition. The identification of a muco-enteric symptom cluster across the whole cohort also raises the suggestion that MIS-C may be the severe end of a spectrum of disease.

## Summary box

### What is already known on this topic

The literature to date has consistently reported that children are less severely affected by SARS-CoV-2 infection than adults. Consequently, there is a less information regarding ethnicity, comorbidities, clinical and laboratory findings in children with SARS-CoV-2. A Multisystem Inflammatory Syndrome in Children and Adolescents (MIS-C) temporally associated with SARS-CoV-2 has been widely reported, however all reports to date arise from retrospective cases series which are vulnerable to recall bias.

### What this study adds

This is a large prospective cohort study of children admitted to hospital with laboratory-confirmed covid-19 and the first with prospective recruitment to examine MIS-C. Severe disease was rare and death exceptionally rare, and then occurred in older children with profound co-morbidity. We find that MIS-C cases have been present for longer than reported.

Ethnicity appears to be a factor both in critical care admission and MIS-C. This cohort has also enabled identification of additional clinical (headache, myalgia, fatigue, sore throat and lymphadenopathy) and laboratory characteristics (including low platelets) which may help to refine the WHO criteria for MIS-C.

## End material

The study protocol is available at http://isaric4c.net/protocols; study registry https://www.isrctn.com/ISRCTN66726260.

## Data Availability

We welcome applications for data and material access through our Independent Data and Material Access Committee (https://isaric4c.net).

The lead author (the manuscript's guarantor) affirms that the manuscript is an honest, accurate, and transparent account of the study being reported; that no important aspects of the study have been omitted; and that any discrepancies from the study as planned (and, if relevant, registered) have been explained.

https://isaric4c.net

https://isaric4c.net/sample_access

## Acknowledgments

This work uses data provided by patients and collected by the NHS as part of their care and support #DataSavesLives. We are extremely grateful to the 2,648 frontline NHS clinical and research staff and volunteer medical students, who collected this data in challenging circumstances; and the generosity of the participants and their families for their individual contributions in these difficult times. We also acknowledge the support of Jeremy J Farrar, Nahoko Shindo, Devika Dixit, Nipunie Rajapakse, Piero Olliaro, Lyndsey Castle, Martha Buckley, Debbie Malden, Katherine Newell, Kwame O’Neill, Emmanuelle Denis, Claire Petersen, Scott Mullaney, Sue MacFarlane, Chris Jones, Nicole Maziere, Katie Bullock, Emily Cass, William Reynolds, Milton Ashworth, Ben Catterall, Louise Cooper, Terry Foster, Paul Matthew Ridley, Anthony Evans, Catherine Hartley, Chris Dunn, D. Sales, Diane Latawiec, Erwan Trochu, Eve Wilcock, Innocent Gerald Asiimwe, Isabel Garcia-Dorival, J. Eunice Zhang, Jack Pilgrim, Jane A Armstrong, Jordan J. Clark, Jordan Thomas, Katharine King, Katie Alexandra Ahmed, Krishanthi S Subramaniam, Lauren Lett, Laurence McEvoy, Libby van Tonder, Lucia Alicia Livoti, Nahida S Miah, Rebecca K. Shears, Rebecca Louise Jensen, Rebekah Penrice-Randal, Robyn Kiy, Samantha Leanne Barlow, Shadia Khandaker, Soeren Metelmann, Tessa Prince, Trevor R Jones, Benjamin Brennan, Agnieska Szemiel, Siddharth Bakshi, Daniella Lefteri, Maria Mancini, Julien Martinez, Angela Elliott, Joyce Mitchell, John McLauchlan, Aislynn Taggart, Oslem Dincarslan, Annette Lake, Claire Petersen, Scott Mullaney and Graham Cooke.

## ISARIC Coronavirus Clinical Characterisation Consortium (ISARIC4C) Investigators

Consortium Lead Investigator J Kenneth Baillie, Chief Investigator Malcolm G Semple, Co-Lead Investigator Peter JM Openshaw. ISARIC Clinical Coordinator Gail Carson. Co-Investigators: Beatrice Alex, Benjamin Bach, Wendy S Barclay, Debby Bogaert, Meera Chand, Graham S Cooke, Annemarie B Docherty, Jake Dunning, Ana da Silva Filipe, Tom Fletcher, Christopher A Green, Ewen M Harrison, Julian A Hiscox, Antonia Ying Wai Ho, Peter W Horby, Samreen Ijaz, Saye Khoo, Paul Klenerman, Andrew Law, Wei Shen Lim, Alexander, J Mentzer, Laura Merson, Alison M Meynert, Mahdad Noursadeghi, Shona C Moore, Massimo Palmarini, William A Paxton, Georgios Pollakis, Nicholas Price, Andrew Rambaut, David L Robertson, Clark D Russell, Vanessa Sancho-Shimizu, Janet T Scott, Louise Sigfrid, Tom Solomon, Shiranee Sriskandan, David Stuart, Charlotte Summers, Richard S Tedder, Emma C Thomson, Ryan S Thwaites, Lance CW Turtle, Maria Zambon. Project Managers Hayley Hardwick, Chloe Donohue, Jane Ewins, Wilna Oosthuyzen, Fiona Griffiths. Data Analysts: Lisa Norman, Riinu Pius, Tom M Drake, Cameron J Fairfield, Stephen Knight, Kenneth A Mclean, Derek Murphy, Catherine A Shaw.

## Data and Information System Manager

Jo Dalton, Michelle Girvan, Egle Saviciute, Stephanie Roberts Janet Harrison, Laura Marsh, Marie Connor. Data integration and presentation: Gary Leeming, Andrew Law, Ross Hendry. Material Management: William Greenhalf, Victoria Shaw, Sarah McDonald. Local Principal Investigators: Kayode Adeniji, Daniel Agranoff, Ken Agwuh, Dhiraj Ail, Ana Alegria, Brian Angus, Abdul Ashish, Dougal Atkinson, Shahedal Bari, Gavin Barlow, Stella Barnass, Nicholas Barrett, Christopher Bassford, David Baxter, Michael Beadsworth, Jolanta Bernatoniene, John Berridge, Nicola Best, Pieter Bothma, David Brealey, Robin Brittain-Long, Naomi Bulteel, Tom Burden, Andrew Burtenshaw, Vikki Caruth, David Chadwick, Duncan Chambler, Nigel Chee, Jenny Child, Srikanth Chukkambotla, Tom Clark, Paul Collini, Graham Cooke, Catherine Cosgrove, Jason Cupitt, Maria-Teresa Cutino-Moguel, Paul Dark, Chris Dawson, Samir Dervisevic, Phil Donnison, Sam Douthwaite, Ingrid DuRand, Ahilanadan Dushianthan, Tristan Dyer, Cariad Evans, Chi Eziefula, Chrisopher Fegan, Adam Finn, Duncan Fullerton, Sanjeev Garg, Sanjeev Garg, Atul Garg, Jo Godden, Arthur Goldsmith, Clive Graham, Elaine Hardy, Stuart Hartshorn, Daniel Harvey, Peter Havalda, Daniel B Hawcutt, Maria Hobrok, Luke Hodgson, Anita Holme, Anil Hormis, Michael Jacobs, Susan Jain, Paul Jennings, Agilan Kaliappan, Vidya Kasipandian, Stephen Kegg, Michael Kelsey, Jason Kendall, Caroline Kerrison, Ian Kerslake, Oliver Koch, Gouri Koduri, George Koshy, Shondipon Laha, Susan Larkin, Tamas Leiner, Patrick Lillie, James Limb, Vanessa Linnett, Jeff Little, Michael MacMahon, Emily MacNaughton, Ravish Mankregod, Huw Masson, Elijah Matovu, Katherine McCullough, Ruth McEwen, Manjula Meda, Gary Mills, Jane Minton, Mariyam Mirfenderesky, Kavya Mohandas, Quen Mok, James Moon, Elinoor Moore, Patrick Morgan, Craig Morris, Katherine Mortimore, Samuel Moses, Mbiye Mpenge, Rohinton Mulla, Michael Murphy, Megan Nagel, Thapas Nagarajan, Mark Nelson, Igor Otahal, Mark Pais, Selva Panchatsharam, Hassan Paraiso, Brij Patel, Justin Pepperell, Mark Peters, Mandeep Phull, Stefania Pintus, Jagtur Singh Pooni, Frank Post, David Price, Rachel Prout, Nikolas Rae, Henrik Reschreiter, Tim Reynolds, Neil Richardson, Mark Roberts, Devender Roberts, Alistair Rose, Guy Rousseau, Brendan Ryan, Taranprit Saluja, Aarti Shah, Prad Shanmuga, Anil Sharma, Anna Shawcross, Jeremy Sizer, Richard Smith, Catherine Snelson, Nick Spittle, Nikki Staines, Tom Stambach, Richard Stewart, Pradeep Subudhi, Tamas Szakmany, Kate Tatham, Jo Thomas, Chris Thompson, Robert Thompson, Ascanio Tridente, Darell Tupper - Carey, Mary Twagira, Andrew Ustianowski, Nick Vallotton, Lisa Vincent-Smith, Shico Visuvanathan, Alan Vuylsteke, Sam Waddy, Rachel Wake, Andrew Walden, Ingeborg Welters, Tony Whitehouse, Paul Whittaker, Ashley Whittington, Meme Wijesinghe, Martin Williams, Lawrence Wilson, Sarah Wilson, Stephen Winchester, Martin Wiselka, Adam Wolverson, Daniel G Wooton, Andrew Workman, Bryan Yates, Peter Young.

## Contributions: Conceptualization

OVS, KAH, LS, PO, JSN-V-T, PWH, LM, GC, WJD, PJMO, JKB, EMH, ABD, MGS. Data curation: SH, MG, LM, EMH, ABD. Analysis: OVS, LPo, CF, TMD, SS, CE, MGP, SL, IPS, EMH, ABD, MGS.

## Funding

JSN-V-T, PWH, GC, PJMO, JKB, ABD, MGS. Investigation: OVS, KAH, LT, LP, SL, EMH, ABD. Methodology: OVS, KAH, LT, LPo, SL, EMH, ABD, MGS. Administration: KAH, LT, HH, SH, MG, CD, LPa, LS, LM. Resources: LS, PO, PWH, LM, MGS. Software: OVS, CF, TMD, SS, CE, LM. Supervision: HH, SL, PO, JSN-V-T, LM, PJMO, JKB, ABD, MGS. Validation: KAH, LT, LPo.

## Visualisation

OVS, CF, TMD, SS, CE, MGP, EMH, ABD. Writing original draft: OVS, KAH, LT, LPo, SL, EMH, ABD, MGS. Writing review and Editing: OVS, KAH, LT, LPo, CF, TMD, MGP, LPa, SL, IPS, JSN-V-T, JKB, EMH, ABD, MGS.

## Funding

This work is supported by grants from: the National Institute for Health Research [award CO-CIN-01], the Medical Research Council [grant MC_PC_19059] and by the National Institute for Health Research Health Protection Research Unit (NIHR HPRU) in Emerging and Zoonotic Infections at University of Liverpool in partnership with Public Health England (PHE), in collaboration with Liverpool School of Tropical Medicine and the University of Oxford [NIHR award 200907], Wellcome Trust and Department for International Development [215091/Z/18/Z], and the Bill and Melinda Gates Foundation [OPP1209135], and Liverpool Experimental Cancer Medicine Centre for providing infrastructure support for this research (Grant Reference: C18616/A25153). JSN-V-T is seconded to the Department of Health and Social Care, England (DHSC). The views expressed are those of the authors and not necessarily those of the DHSC, DID, NIHR, MRC, Wellcome Trust or PHE.

### Competing interests

All authors have completed the ICMJE uniform disclosure form at www.icmje.org/coi_disclosure.pdf and declare: JSN-V-T reports grants from Department of Health and Social Care, England, during the conduct of the study; PWH reports grants from Wellcome Trust / Department for International Development / Bill and Melinda Gates Foundation, grants from NIHR, during the conduct of the study; PJMO reports personal fees from Consultancy, grants from MRC, grants from EU Grant, grants from NIHR Biomedical Research Centre, grants from MRC/GSK, grants from Wellcome Trust, grants from NIHR (HPRU), grants from NIHR Senior Investigator, personal fees from European Respiratory Society, grants from MRC Global Challenge Research Fund, outside the submitted work; and The role of President of the British Society for Immunology was an unpaid appointment but my travel and accommodation at some meetings is provided by the Society; AMD reports grants from Department of Health and Social Care, during the conduct of the study; grants from Wellcome Trust, outside the submitted work; JKB reports grants from DHSC National Institute of Health Research UK, grants from Medical Research Council UK, grants from Wellcome Trust, grants from Fiona Elizabeth Agnew Trust, grants from Intensive Care Society, grants from Chief Scientist Office, during the conduct of the study; MGS reports grants from DHSC National Institute of Health Research UK, grants from Medical Research Council UK, grants from Health Protection Research Unit in Emerging & Zoonotic Infections, University of Liverpool, during the conduct of the study; other from Integrum Scientific LLC, Greensboro, NC, USA, outside the submitted work; the remaining authors declare no competing interests; no financial relationships with any organisations that might have an interest in the submitted work in the previous three years; and no other relationships or activities that could appear to have influenced the submitted work.

### Ethical approval

Ethical approval was given by the South Central - Oxford C Research Ethics Committee in England (Ref 13/SC/0149), the Scotland A Research Ethics Committee (Ref 20/SS/0028), and the WHO Ethics Review Committee (RPC571 and RPC572, 25 April 2013).

### Data sharing

We welcome applications for data and material access through our Independent Data and Material Access Committee (https://isaric4c.net). The lead author (the manuscript’s guarantor) affirms that the manuscript is an honest, accurate, and transparent account of the study being reported; that no important aspects of the study have been omitted; and that any discrepancies from the study as planned (and, if relevant, registered) have been explained.

### Dissemination to participants and related patient and public communities

ISARIC4C has a public facing website and twitter account @CCPUKstudy. We are engaging with print and internet press, television, radio, news, and documentary programme makers. We will explore distribution of findings with The Asthma UK and British Lung Foundation Partnership, and take advice from NIHR Involve and GenerationR Alliance Young People’s Advisory Groups.

The Corresponding Author has the right to grant on behalf of all authors and does grant on behalf of all authors, a worldwide licence (http://www.bmj.com/sites/default/files/BMJ%20Author%20Licence%20March%202013.doc) to the Publishers and its licensees in perpetuity, in all forms, formats and media (whether known now or created in the future), to i) publish, reproduce, distribute, display and store the Contribution, ii) translate the Contribution into other languages, create adaptations, reprints, include within collections and create summaries, extracts and/or, abstracts of the Contribution and convert or allow conversion into any format including without limitation audio, iii) create any other derivative work(s) based in whole or part on the on the Contribution, iv) to exploit all subsidiary rights to exploit all subsidiary rights that currently exist or as may exist in the future in the Contribution, v) the inclusion of electronic links from the Contribution to third party material where-ever it may be located; and, vi) licence any third party to do any or all of the above. All research articles will be made available on an open access basis (with authors being asked to pay an open access fee—see http://www.bmj.com/about-bmj/resources-authors/forms-policies-and-checklists/copyright-open-access-and-permission-reuse). The terms of such open access shall be governed by a Creative Commons licence—details as to which Creative Commons licence will apply to the research article are set out in our worldwide licence referred to above.

## Notes

### Clinical Trial

ISRCTN66726260

### Clinical Protocols

https://isaric4c.net/protocols/

